# Sex differences in ACE2, TMPRSS2, and HLA-DQA2 expression in gray matter: Implications for post-COVID-19 neurological symptoms

**DOI:** 10.1101/2024.11.04.24316706

**Authors:** Shelli R. Kesler, Alexa De La Torre Schutz, Oscar Y. Franco Rocha, Kimberly Lewis

## Abstract

COVID-19 has been associated with sex differences in terms of mortality and morbidity. Viral entry proteins including those regulated by ACE2 and TMPRSS2 may play a role, but few studies have been conducted to date and none have examined sex differences in brain expression. Additionally, HLA-DQA2 expression has emerged as a potential moderator of COVID-19 outcomes. Using non-invasive imaging transcriptomics, we measured ACE2, TMPRSS2, and HLA-DQA2 mRNA expression in gray matter volumes using MRI scans obtained from 1,045 healthy adults aged 21-35 years (44% male) imaged prior to the COVID-19 pandemic. ACE2 (t = 9.24, p < 0.001, d = 0.576), TMPRSS2 (t = 24.66, p < 0.001, d = 1.54), and HLA-DQA2 (t = 3.70, p < 0.001, d = 0.231) expression was significantly higher in males compared to females. Bayesian network analysis indicated significant (p < 0.05) positive causal paths from ACE2 to HLA-DQA2 (B = 0.282), ACE2 to TMPRSS2 (B = 0.357), and TMPRSS2 to HLA-DQA1 (B = 0.139) and a negative causal path from sex (males = -1, females = 1) to TMPRSS2 (B = -0.607). Our results have important implications for neurological symptoms associated with COVID-19 and long COVID including complex interactions between viral entry proteins and immune responses, sex-related disparities in symptom reporting and diagnosis, assessment of neurological problems after COVID-19, and potential COVID-19 related syndemics. However, further research is needed to determine gene expression patterns by sex and COVID-19 outcomes, to evaluate additional genes that may influence neurologic status, and studies that include objective assessments of neurologic outcomes.

## Introduction

COVID-19 mortality and morbidity are higher in men compared to women (1-4). The mechanisms underlying this disparity remain unclear but existing evidence suggests that sex differences in expression of ACE2 and TMPRSS2 may play a significant role. SARS-CoV-2, the virus responsible for COVID-19, uses ACE2 receptors to enter human cells (5). TMPRSS2 is crucial for facilitating the entry of some viruses, including SARS-CoV-2, into host cells by cleaving viral spike proteins (5-7).

HLA-DQA2 expression may moderate the duration of COVID-19 infection (8). HLA-DQA2 is a gene that encodes part of the major histocompatibility complex (MHC) class II molecule. The primary function of HLA-DQA2, like other MHC class II molecules, is to present antigens to CD4+ T helper cells, initiating immune responses against pathogens. Sex differences in HLA-DQA2 expression have not been extensively studied but it is well-known that sex can influence immune function.

Although the WHO declared that COVID-19 is no longer a global health emergency, many individuals experience persistent symptoms post-infection. These symptoms are known as long COVID, or Post-acute Sequelae of SARS-CoV-2 Infection (PASC) (9, 10). Neurological symptoms, including cognitive dysfunction, are common characteristics of PASC. In fact, “brain fog” is a primary symptom of PASC, affecting an estimated 20% of patients (11). Our group and others have shown significant, widespread brain changes months after COVID-19 infection, even in mild to moderate cases (12-15). Although COVID-19 mortality tends to be worse in men, our group and others have observed that PASC, including cognitive and other neurological symptoms, may be more prevalent in women (16-18).

ACE2 expression has been shown to be dysregulated in the brain after COVID-19, especially in patients with significant neurologic symptoms (19). The specific functions and roles of TMPRSS in the brain and cognition are less well-documented. However, a recent study provided evidence that ACE2 and TMPRSS2 genes are overexpressed in similar regions throughout the brain, including orbitofrontal cortex (20). MHC class II molecules, including those encoded by HLA-DQA2, can be expressed in the brain under conditions of neuroinflammation. There is emerging evidence that MHC class II molecules may contribute to the development or exacerbation of neurodegenerative diseases, such as multiple sclerosis and Alzheimer’s disease, by modulating immune responses in the brain (21, 22).

One study showed that ACE2 and/or TMPRSS2 expression in various tissues did not differ between males and females (23), but did not compare expression levels in the brain. Another showed that ACE2 expression is higher in males and increases more with age compared to females (24) but this was based on peripheral analysis so again, did not measure levels in the brain. It is also unclear whether there are sex differences in HLA-DQA2 expression. Understanding how genes associated with COVID-19 are expressed in individuals who were never exposed to the virus provides a baseline for comparison. This is important for distinguishing the natural variability in gene expression from potential alterations caused by the virus itself. We aimed to measure the mRNA expression of ACE2, TMPRSS2, and HLA-DQA2 in brain tissue. However, most individuals have had a COVID-19 infection at this point.

Therefore, we applied non-invasive imaging transcriptomics methods to a large, publicly available dataset of healthy adults who underwent brain MRI prior to the COVID-19 pandemic. We hypothesized that mRNA expression levels would differ between males and females.

## Methods

### Participants

We obtained retrospective T1-weighted brain MRI scans and basic demographic data from Harvard University Brain Genomics Superstruct Project (GSP) open access data release (25, 26). GSP evaluated 1,570 healthy adults aged 18-35 between 2008 and 2012. We focused on datasets from individuals who were 21 years or older to be informative for our ongoing studies of PASC-related cognitive dysfunction in younger adults and thus our sample size for this study was 1,045 (median age = 23 years, IQR = 2, 44% male). Sex was defined as biological sex designated at birth. Participants provided written informed consent in accordance with guidelines established by the Partners Health Care Institutional Review Board and the Harvard University Committee on the Use of Human Subjects in Research and only those who agreed to data sharing were included in the data release. Privacy rights of participants were observed and no personally identifiable information was obtained.

### MRI Data

Brain MRI acquisition details are described elsewhere (26). Briefly, imaging data were collected using 3 Tesla Siemens Tim Trio scanners (Siemens Healthcare, Erlangen, Germany) at Harvard University and Massachusetts General Hospital with the vendor-supplied 12-channel head coil. T1-weighted anatomic MRI data were acquired using a high-resolution (1.2mm isotropic) multi-echo magnetization-prepared gradient-echo image in the sagittal orientation (2.20 minute scan duration).

### Imaging Transcriptomics

Gray matter volumes were segmented from MRI with Voxel-Based Morphometry in Statistical Parametric Mapping v12 (Wellcome Trust Centre for Neuroimaging, London, UK) in Matlab v2023b (Mathworks, Inc., Natick, MA). We employed Diffeomorphic Anatomical Registration Through Exponentiated Lie Algebra (DARTEL), which uses a large deformation framework to preserve topology and employs customized, sample-specific templates resulting in superior image registration. Successful normalization was confirmed using visual and quantitative quality assurance methods.

We obtained brain transcriptome data for ACE2, TMPRSS2, and HLA-DQA2 from the Allen Human Brain Atlas (AHBA), which is currently considered the most comprehensive transcriptional brain map available (27). AHBA was developed using six adult human donor brains to provide expression data from tens of thousands of genes measured from thousands of brain regions (28). We used the Multimodal Environment for Neuroimaging and Genomic Analysis (MENGA) toolbox v3.1 in Matlab v2023b to correlate the gray matter volume maps of each participant with patterns of AHBA expression from the 3 selected genes. Imaging data for each participant were resampled into AHBA coordinates with a 5mm resolution. The expression data for each AHBA donor, for each gene was sampled from each of 169 AHBA regions resulting in a 169×3 region-by-gene expression matrix for each gene. Principal Components Analysis was performed on the region-by-gene expression matrix to identify components explaining at least 95% variance across the AHBA donors. The component scores were then entered as independent variables into a weighted, multiple linear regression analysis with the corresponding gray matter volume values (169×1 vector) from the participant as the dependent variable. This resulted in an R squared value for each gene for each participant representing the mRNA expression in total gray matter.

### Reliability of Genomic Data

Most of the genes included in AHBA have multiple probes with some showing more reliable expression patterns than others. First, the current accuracy of the probe to gene mapping was verified and probe data were normalized to z-scores. Representative probes were then selected in a data-driven manner considering between-donor homogeneity and the distributions of probe data. Genomic expression variability between donors was evaluated using an intraclass correlation coefficient (ICC). Most of the tissue samples used to develop AHBA were obtained only from the left hemisphere and therefore, we employed a binary mask to limit our analysis to the left hemisphere (29).

### Statistical Analyses

mRNA expression in gray matter was compared between groups using t-tests. We conducted a two stage path analysis to determine the causal relationships for mRNA expression. Given that little is known regarding these relationships for our selected genes, we first conducted an exploratory Bayesian network analysis. This method learns the conditional dependencies between variables without any prior assumptions. We utilized a hill-climbing approach by optimizing the Bayesian information criterion to identify the best fit network. The conditional independence/dependence relationships among variables were encoded with a directed acyclic graph. We then supplemented this with structural equation modeling (SEM) to provide the more common model statistics such as RMSEA using the Bayesian network results as the model framework.

Statistical analyses were conducted using the R Statistical Package (v4.4.1) including the “bnlearn” library for Bayesian network analysis and the “lavaan” library for SEM. Alpha level for all analyses was p < 0.05 and multiple comparisons were corrected using the false discovery rate (FDR). Effect sizes for t-tests were computed using Cohen’s d.

## Results

As shown in Table 1 and Figure 1, ACE2 (t = 9.24, p < 0.001, d = 0.576), TMPRSS2 (t = 24.66, p < 0.001, d = 1.54), and HLA-DQA2 (t = 3.70, p < 0.001, d = 0.231) expression was significantly higher in males compared to females.

**Table 1.**
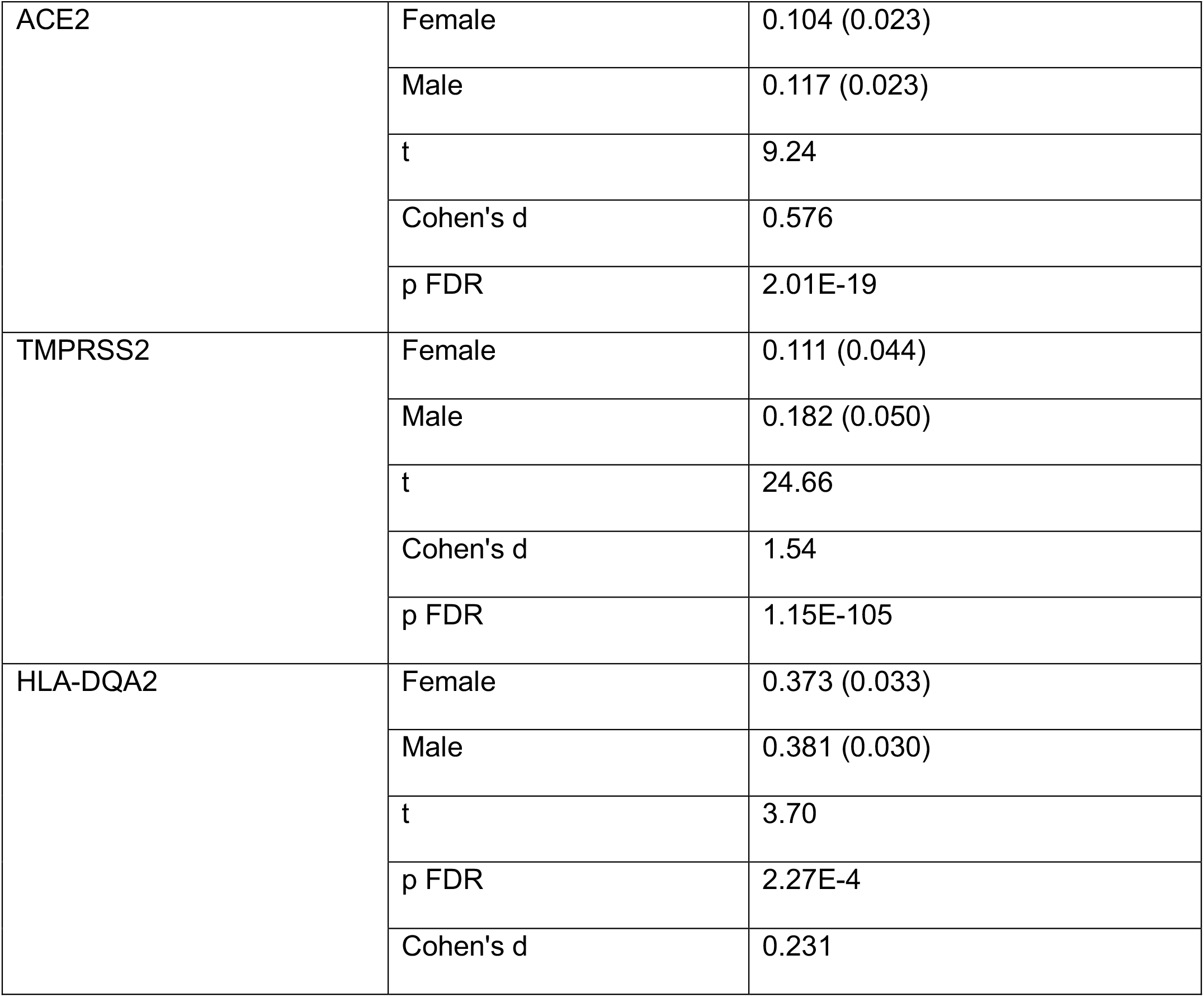
Genomic expression in gray matter for females (N = 591) and males (N = 454). Data are shown as mean (standard deviation). Data values are the R squared statistic for the relationship between mRNA expression and gray matter volume. The false discovery rate (FDR) corrected p values are presented.

|  |  |  |
| --- | --- | --- |
| ACE2 | Female | 0.104 (0.023) |
|  | Male | 0.117 (0.023) |
|  | t | 9.24 |
|  | Cohen's d | 0.576 |
|  | p FDR | 2.01E-19 |
| TMPRSS2 | Female | 0.111 (0.044) |
|  | Male | 0.182 (0.050) |
|  | t | 24.66 |
|  | Cohen's d | 1.54 |
|  | p FDR | 1.15E-105 |
| HLA-DQA2 | Female | 0.373 (0.033) |
|  | Male | 0.381 (0.030) |
|  | t | 3.70 |
|  | p FDR | 2.27E-4 |
|  | Cohen's d | 0.231 |

**Figure 1.**
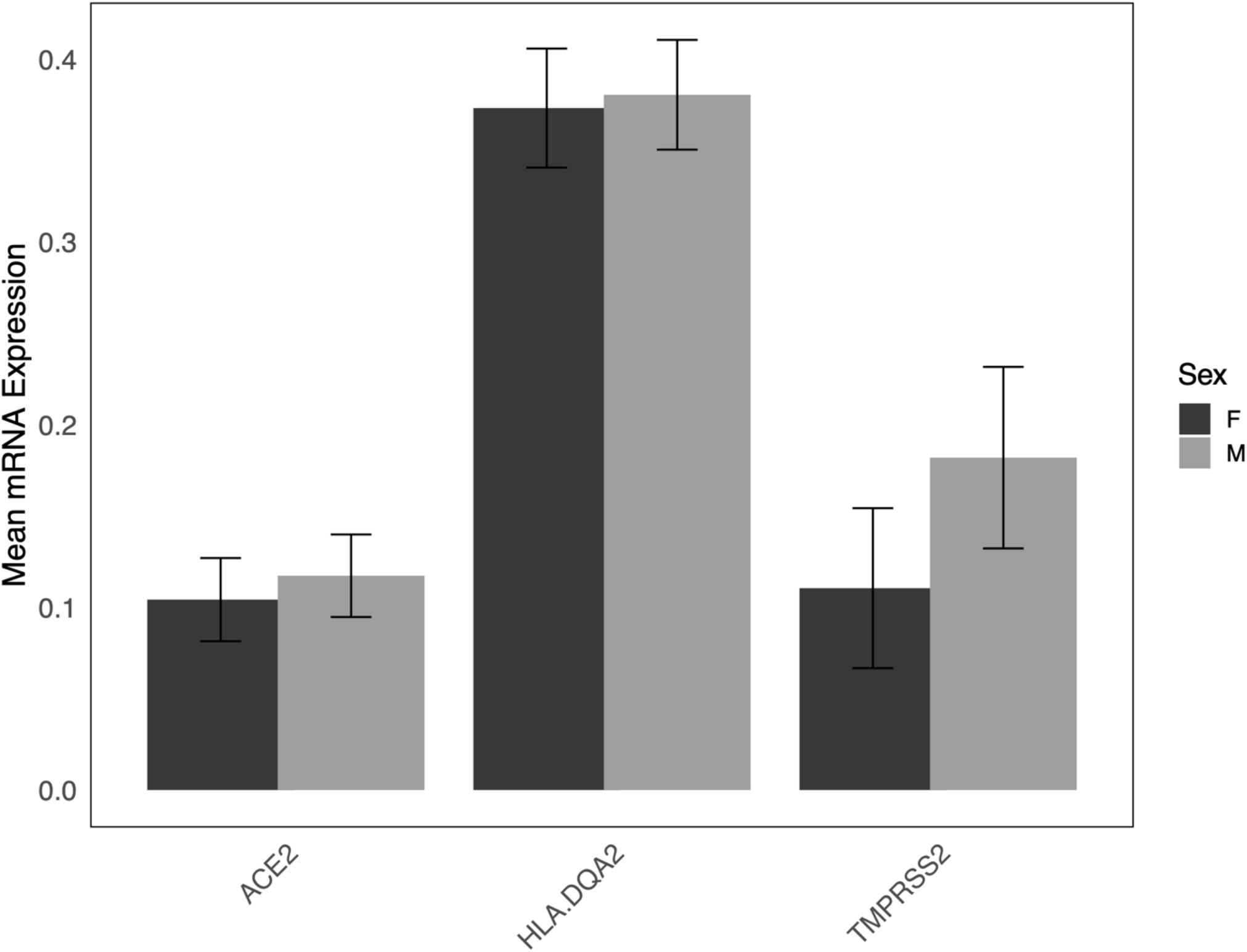
Sex differences in ACE2, TMPRSS2, and HLA-DQA2 mRNA expression in gray matter. Bar graphs indicate mean mRNA expression for females (F) and males (M). Error bars represent the standard deviation. All comparisons were significant (p < 0.001, false discovery rate corrected).

Bayesian network analysis indicated a significant directed acyclic graph as illustrated in Figure 2 with positive causal paths from ACE2 to HLA-DQA2 (B = 0.282), ACE2 to TMPRSS2 (B = 0.357), and TMPRSS2 to HLA-DQA1 (B = 0.139) and a negative causal path from sex (males = -1, females = 1) to TMPRSS2 (B = -0.607). Based on this framework, a supplementary SEM (Figure 2) demonstrated fit statistics of RMSEA = 0.00 (95% CI = 0.00 to 0.00), CFI = 1.00, and TLI = 1.00).

**Figure 2.**
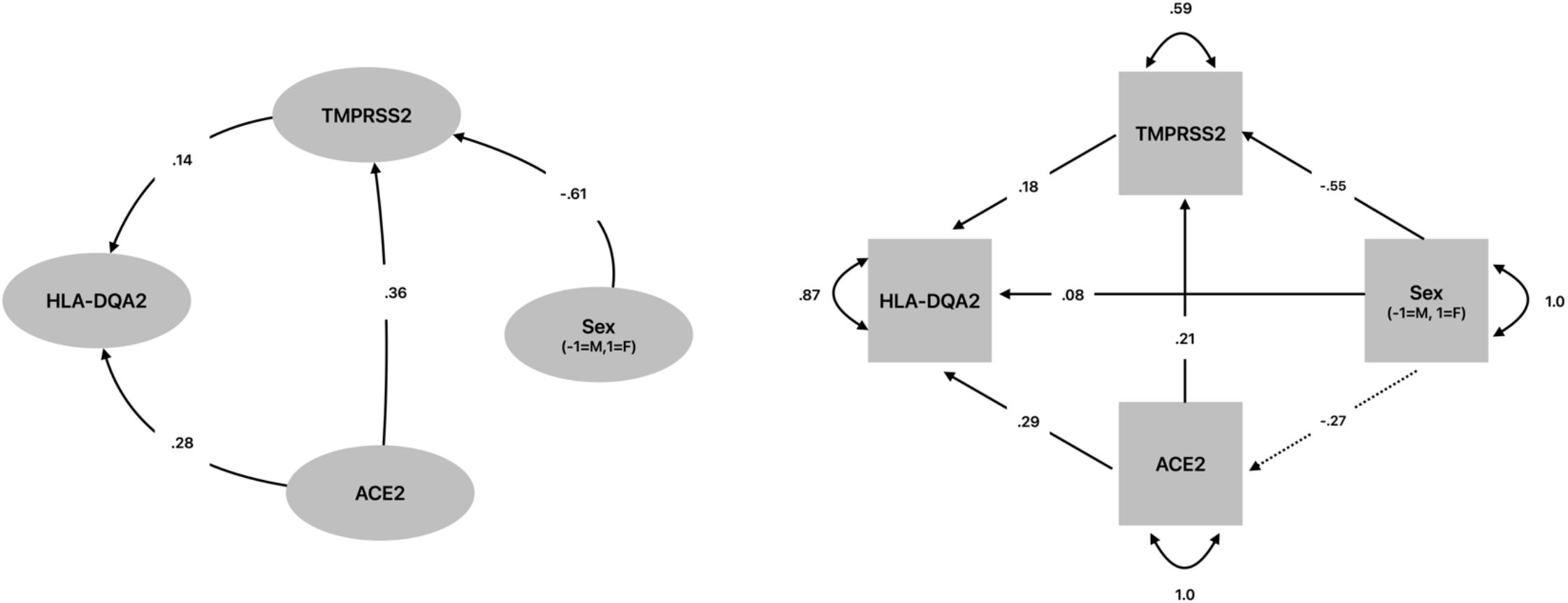
Causal relationships among ACE2, TMPRSS2, and HLA-DQA2 mRNA expression in gray matter determined by Bayesian network analysis directed acyclic graph (left) and subsequent structural equation modeling (right) where RMSEA = 0.00 (95%CI = 0.00 to 0.00), CFI = 1.00, and TLI = 1.0). Standardized coefficients are presented.

The ICC’s across AHBA donors were 0.450 (SD = 0.219) for ACE2, 0.378 (SD = 0.195) for TMPRSS2, and 0.208 (SD = 0.151) for HLA-DQA2.

## Discussion

Using imaging transcriptomics, we demonstrated that the expression of genes associated with COVID-19 infection in gray matter, including ACE2, TMPRSS2, and HLA-DQA2, is significantly higher in men compared to women. There was a large effect for TMPRSS2 (d = 1.5), a medium effect for ACE2 (d = 0.58), and a small effect for HLA-DQA2 (d = 0.23). These findings suggest a potential mechanism for sex differences in COVID-related outcomes, particularly in terms of neurological symptoms.

Our findings are consistent with previous studies examining non-brain tissues which suggest that ACE2 expression is generally higher in men than in women (3). Higher ACE2 expression in tissues such as lung for example, help explain the increased COVID-19 mortality in men. Few studies have examined sex differences in TMPRSS2 or HLA-DQA2. One study found no sex differences in TMPRSS2 expression within tissue from select organs involved in gastrointestinal, respiratory, urinary, and endocrine function (23). However, expression in brain tissue was not compared between men and women.

Higher ACE2 and TMPRSS2 expression in the gray matter of males in the context of higher incidence of PASC-related neurological symptoms in women requires further discussion. There are several potential explanations including immune response differences. Females tend to have stronger immune responses which is beneficial for fighting diseases but can result in higher inflammation (30). Women also tend to be more vulnerable to certain autoimmune disorders, including those that affect brain structure and function (31). Our path analysis results suggested that ACE2 and TMPRSS2 upregulate HLA-DQA2, which is critically involved in immune response. Thus upregulation of an already heightened microglial response in the brain may occur in women despite lower COVID-19 related gene expression overall.

Alternatively, symptom reporting and detection bias may play a role. Women are more likely to report certain symptoms and healthcare providers tend to be more likely to diagnosis certain symptoms in women compared to men, even with equivocal symptom presentations (32-36). These include neurologic and neuropsychiatric symptoms such as fatigue, depression, anxiety, and cognitive dysfunction. Most studies focused on PASC-related neurological effects have employed subjective measures. While these can be more sensitive to underlying changes in neurologic function, they are not interchangeable with objective assessments (37).

In terms of objective cognitive testing for example, some studies demonstrated lower objective cognitive function in females after COVID-19 (38, 39) while others showed no sex differences. (40-43) However, these studies used different assessments and had various sample characteristics. Importantly, most studies have not examined sex effects. For example, we identified 53 studies of PASC-related cognitive dysfunction (Supplementary Materials) and found that only the 6 mentioned above (11%) investigated sex differences in objective cognitive tests. Given the over-reliance on subjective measures, which will bias symptom incidence towards women, and the general lack of studies regarding sex differences, it remains unclear whether or not women truly have a larger vulnerability to PASC-related neurological symptoms.

Quantitative neuroimaging is another objective measure of neurologic status. Using these methods, we showed that men have a 99% higher risk of accelerated brain aging after COVID-19 compared to women despite there being no significant differences in objective cognitive testing performance (12). Another study demonstrated reduced functional activity in a prefrontal cortex as well as lower objective cognitive testing performance in women compared to men after COVID-19 (39). However, as noted above, we could find few studies that examined sex differences and therefore, further research is needed.

HLA-DQA2 showed the highest expression in gray matter, accounting for 37% of the variance in females and 38% of the variance in males, on average. There was a significant but small effect for HLA-DQA2 being higher in men compared to women and our path analysis suggested that both ACE2 and TMPRSS2 upregulated HLA-DQA2 after controlling for sex. Based on research indicating that increased expression of HLA-DQA2 may be associated with shorter COVID-19 infection duration (8), its expression in gray matter may help reduce neurologic effects. However, HLA-DQA2 and other MHC Class II molecules (e.g., HLA-DRB1) are involved in neurodegenerative conditions such as Alzheimer’s disease, multiple sclerosis, and Parkinson’s disease (22, 44, 45), which suggests a complex relationship. Many biological systems, including the brain, operate in states of critical balance where deviations—whether excess or deficiency—can lead to dysfunction. For example, ACE2 expression is associated with neuroprotective effects, yet several studies have noted elevated ACE2 levels in patients with Alzheimer’s disease (46-48). COVID-19 infection may disrupt these systems resulting in neurological resilience or vulnerability, despite baseline gene expression levels. Accordingly, our present findings involved healthy adults who were never exposed to COVID-19. Viral effects, such as cytokine storm, may interact with ACE2, TMPRSS2, and HLA-DQA2 brain expression differently in women compared to men. Few studies have been conducted to date and none have compared gene expression patterns by sex and COVID-19 outcomes (3).

Our findings may suggest a potential mechanism for COVID-19 related syndemics. A syndemic, or synergistic epidemic, refers to the simultaneous or sequential occurrence of two or more epidemics or disease clusters within a population, which interact in a way that exacerbates the overall burden of disease and impacts prognosis. Prior studies have shown that COVID-19 infection may worsen existing neurologic conditions such as Alzheimer’s disease and multiple sclerosis (49, 50). Our results suggest that this may occur in part by upregulation of HLA-DQA2 expression in brain tissue by ACE2 and TMPRSS2 within an already pathological state.

Our study provides novel insights regarding the potential contributions of COVID-19 related gene expression to brain structure. However, several limitations should be considered including our lack of data regarding the effects of the virus itself, as mentioned above. Additionally, the ICC’s across AHBA donors for these genes were small to medium indicating heterogeneity in expression. Our path analysis was a very simplistic representation of causal relationships given that there are many other genes involved in gray matter structure and likely in COVID-19 outcomes as well. Our analysis focused on total gray matter volume but there may be regional effects that require further research. Robust contrast maps indicating brain regions that are affected in large samples of COVID-19 survivors with neurological symptoms versus those without symptoms are required. The age range of our sample was restricted by the data available and thus further research is required, especially in older populations who are more vulnerable to COVID-19 related morbidities.

In conclusion, this study uniquely demonstrated sex differences in ACE2, TMPRSS2, and HLA-DQA2 mRNA expression in gray matter volumes of healthy, young adults. Our findings have potential implications for neurological effects of COVID-19 as well as COVID-19 related syndemics that are characterized by neurodegeneration. TMPRSS2 had the largest effect and is thus a strong candidate for future studies regarding the mechanisms of COVID-19 related neurological morbidity. Given our findings of upregulation of HLA-DQA2 by ACE2 and TMPRSS2, the inclusion of other MHC Class II molecules, such as HLA-DRB1, will be important in future studies.

## Data Availability

All data are available via the Harvard University Brain Genomics Superstruct Project (GSP) open access project (https://dataverse.harvard.edu/dataverse/GSP).

## Acknowledgements

Data were provided by the Brain Genomics Superstruct Project of Harvard University and the Massachusetts General Hospital, (Principal Investigators: Randy Buckner, Joshua Roffman, and Jordan Smoller), with support from the Center for Brain Science Neuroinformatics Research Group, the Athinoula A. Martinos Center for Biomedical Imaging, and the Center for Human Genetic Research. Twenty individual investigators at Harvard and MGH generously contributed data to the overall project.

